# Unawareness for chronic kidney disease is high in all stages, age groups and comorbidities – and higher in women than in men

**DOI:** 10.1101/2021.05.21.21257582

**Authors:** Susanne Stolpe, Bernd Kowall, Christian Scholz, Andreas Stang, Cornelia Blume

## Abstract

**Background:** Chronic kidney disease (CKD) is associated with an increased risk for cardiovascular events, hospitalizations or mortality. In populations aged ≥40 years, CKD is as prevalent as diabetes or coronary heart disease. Awareness for CKD though is generally low in public, patients and physicians, which hinders early diagnosis and treatment to decelerate disease progress.

**Method:** We analyzed baseline data collected in 2010 from 3,334 participants with CKD stages 1-5 from German CKD cohorts and registries. CKD unawareness and 95%-confidence intervals (CI) was estimated according to patients’ answer to the question whether they had ever been told to suffer from a CKD. Prevalence ratios (PR) with 95%-CI were estimated in categories of age, sex, CKD stages, BMI, hypertension, diabetes and other relevant comorbidities.

**Results:** CKD unawareness was high, reaching 82% (95% CI: 80%-84%) for CKD stages 1 or 2, 71% (68%-73%) in CKD 3a, 49% (45%-54%) in CKD 3b and still 30% (24%-36%) in CKD4, in each stage increasing with age. CKD unawareness was similarly high in patients with hypertension, diabetes or cardiovascular comorbidities. Women were more often unaware than men (PR=1.07 (1.02;1.12)), this sex difference increased with increasing CKD stage. Macroalbuminuria (PR=0.90 (0.82; 1.00)), anemia (PR=0.78 (0.73; 0.83)) and BMI ≥40 (PR=0.88 (0.77; 1.00)) were associated with higher CKD awareness.

**Conclusion:** Even in older patients or in patients with comorbidities, CKD unawareness was high. Sex differences were largest in later stages. Guideline oriented treatment of patients with hypertension or diabetes could increase awareness. Patient-physician communication about CKD might be amendable.

## Background

Chronic kidney disease (CKD) is a highly prevalent disease which is widely unrecognized in its relevance for personal health consequences and its impact on societal health systems’ spending (1-3). CKD remains asymptomatic throughout a long time until reaching later stages defined by a severely reduced renal function. CKD is associated with a higher risk for renal failure and other non-renal health outcomes such as cardiovascular diseases, hospitalizations and premature mortality (1). Discussions in the medical community whether CKD in elderly should be regarded as normal physiologic aging process or be labelled as ‘disease’ are still ongoing (4). In a population aged 40 years and older, prevalence of CKD is about 10% and comparable to the prevalence of other chronic diseases as diabetes, depression, or coronary heart disease (2, 5-7). Contrasting to these, for CKD a high public and patient unawareness has been reported: about 80% in early stages, and about 30% in later stages in populations from USA, Australia or Taiwan (8), even in patients with markers for renal dysfunction (9). In Germany, 72% of CKD patients from a general population did not know about their disease (10), unawareness was even high in patients hospitalized due to cardiovascular diseases (11). Public knowledge about CKD is scarce (12). Screening for CKD even in populations at higher risk is mostly lacking – resulting in late diagnosis and treatment. Although the estimated glomerular filtration rate (eGFR) – an estimation of renal function based on creatinine in serum – is calculated and printed automatically on patients’ laboratory reports, this seems not to lead to a routinely ascertainment of kidney function and diagnosis of CKD. Communicating information about CKD and its relevance for health to patients seems to be more difficult in CKD than in other chronic diseases with negative impact on patients’ involvement and compliance to treatment (13, 14).

CKD unawareness can result from either not yet being diagnosed or not being told about by a physician or not fully grasp the meaning of CKD information.

We wanted to estimate CKD unawareness focusing on patients with CKD related risk factors, as these patients can be expected to be targeted in primary health care for monitoring renal function according to guidelines. Among these patients, we wanted to identify demographic and clinical factors that are associated with low CKD awareness.

## Material and Methods

We used baseline data collected in 2010 from German CKD research cohorts and registries that have been compiled according to a standardized protocol into a joint database –the “CORE database”, see supplementary file. From this database, we included participants from the Diabetes Cohort (DIACORE), the Coronary Artery Disease – Renal Failure Registry (CAD-REF) and the Berlin Initiative Study (BIS)) (N= 8,421). Participants in these cohort studies either had angiographically documented coronary artery disease (CAD-REF), self-reported diabetes (DIACORE) or were of higher age (BIS)(15-17). Baseline data include age, sex and clinical information such as blood pressure and BMI, questionnaire data about history of comorbidities and medications as well as centrally analyzed laboratory data.

For estimation of CKD unawareness, we included only those participants (N=3,334) who had a prevalent CKD stages 1-5 according to the KDIGO (Kidney Disease Improving Global Outcomes (18)) guideline definition using laboratory information on eGFR and albuminuria. Patients with CKD stage 5 were not included in the analyses of CKD unawareness stratified by CKD stage, as only one patient out of 29 was unaware.

### Variable definition

Unknown CKD was defined according to the patients’ answer to the question ‘Have you ever been told, that you have kidney disease or kidney stones?’. Laboratory data was centrally analyzed by SynLab, Germany. Serum creatinine was measured using the enzymatic method on a Roche Cobas 6000 C502, eGFR (ml/min/1.73m^2^) was calculated using the CKD-Epi equation (19).

We categorized age in age groups <50, 50-59, 60-69, 70-79 and ≥80 years.

Intake of anti-hypertensive medication was coded according to the patients’ answer to the respective question. Intake of anti-diabetic medication was coded if participants reported taking medications with an ATC-code starting with A10, indicating anti-diabetic drugs. HbA1c had been measured in 2,471 patients.

An albumin/creatinine ratio (ACR) ≥30 indicated a micro-, an ACR ≥300 a macroalbuminuria. BMI was categorized in <25, 25-<30, 30-<40 and ≥40kg/m^2^. The KDIGO guideline 2012 recommends blood pressure lowering treatment in CKD patients with a blood pressure ≥140mmHg systolic or ≥90mmHg diastolic (18). We used this threshold to define good and less good blood pressure control. We used the threshold of ≥160mmHg systolic or ≥95mmHg diastolic to indicate inadequate blood pressure control. KDIGO defines a HbA1c of <7% as optimal glucose control and recommends a target HbA1c of ∼ 7% in CKD patients (18). We categorized HbA1c in <7, 7-<7.5% 7.5-<9% and >= 9% representing optimal, adequate, less well controlled and, uncontrolled glucose.

Cardiovascular disease was coded as prevalent, if a patient positively answered a question about the history of myocardial infarction, bypass surgery, stroke or heart failure.

For each participant, we estimated the probability of reaching end stage renal disease (ESRD) within the next five years using the Kidney-failure-risk-equation (KFRE). KFRE includes the variables age, sex, serum creatinine and ACR. (20)

We calculated a sum score of comorbidities and unfavorable conditions in relation to CKD to mirror participants’ burden of CKD risk factors. The score included albuminuria, anti-hypertensive medication, anti-diabetic medication, history of cardiovascular disease, anemia, obesity and ≥70 years.

### Statistical methods

We estimated the prevalence of CKD unawareness and 95% exact confidence intervals (CI) in all patients, and stratified by age, sex, CKD stage, categories of BMI and blood pressure, anti-hypertensive and anti-diabetic medication, and comorbidities such as anemia, albuminuria, stroke, myocardial infarction or heart failure. In patients taking anti-diabetic medication, we calculated the prevalence of CKD unawareness by category of glucose control. We selected insulin, metformin and metformin-combined drugs for subgroup analysis of diabetic patients, as intake of insulin indicates a more severe diabetes and as metformin is contra-indicated with CKD stage 4. In patients with anti-hypertensive medication, CKD unawareness according to blood pressure categories was calculated. For all selected variables, prevalence ratios (PR) for CKD awareness with 95%-CI were estimated using univariate log-binomial regression models. With categorical variables, the category representing the lowest exposure was used as reference. As we want to describe associations between CKD unawareness and patients’ characteristics to depict the situation in clinical practice, adjustment for confounders is not necessary as stated by Hernan: “if the goal of the observational analysis is purely associational, no adjustment for confounding is necessary” (21). As the size of presented crude PRs and their relevance may be questioned, we nonetheless present sex and age adjusted PRs as well. We did not impute missing values.

All analyses were done in SAS 9.4.

## Results

### Characteristics of participants

Among 3,334 participants, 2,268 (=68%, 95%-CI: 66%-70%) were unaware of their CKD (Tab. 1). 1,219 (37%) patients were classified as CKD stage 1 or 2. In 1,284 (39%) patients a CKD stage 3a and in 611 (18%) CKD stage 3b was present. 220 (7%) had an even more severely decreased renal function. Mean age of the participants was 73.5 (10.4 standard deviation) years, 2.5% were younger than 50 years. Participants had a high prevalence of CKD related risk factors. Proteinuria was prevalent in 2,018 (62%) patients, 2,967 (89%) reported intake of anti-hypertensive medication. Antidiabetic medication was taken by 1,505 patients (45%) and obesity was registered in 1,414 (43%) patients. Participants who were unaware of their CKD differed from those who knew about their disease in prevalence of macroalbuminuria (8.8% vs 13.6%), anemia (20.1% vs 33.6%) or cardiovascular comorbidities (40.5% vs 48%) (Tab. 1). Median eGFR (ml/min/1.73m^2^) was higher in the unaware participants (58.4 vs 47.9).

**Tab 1.**
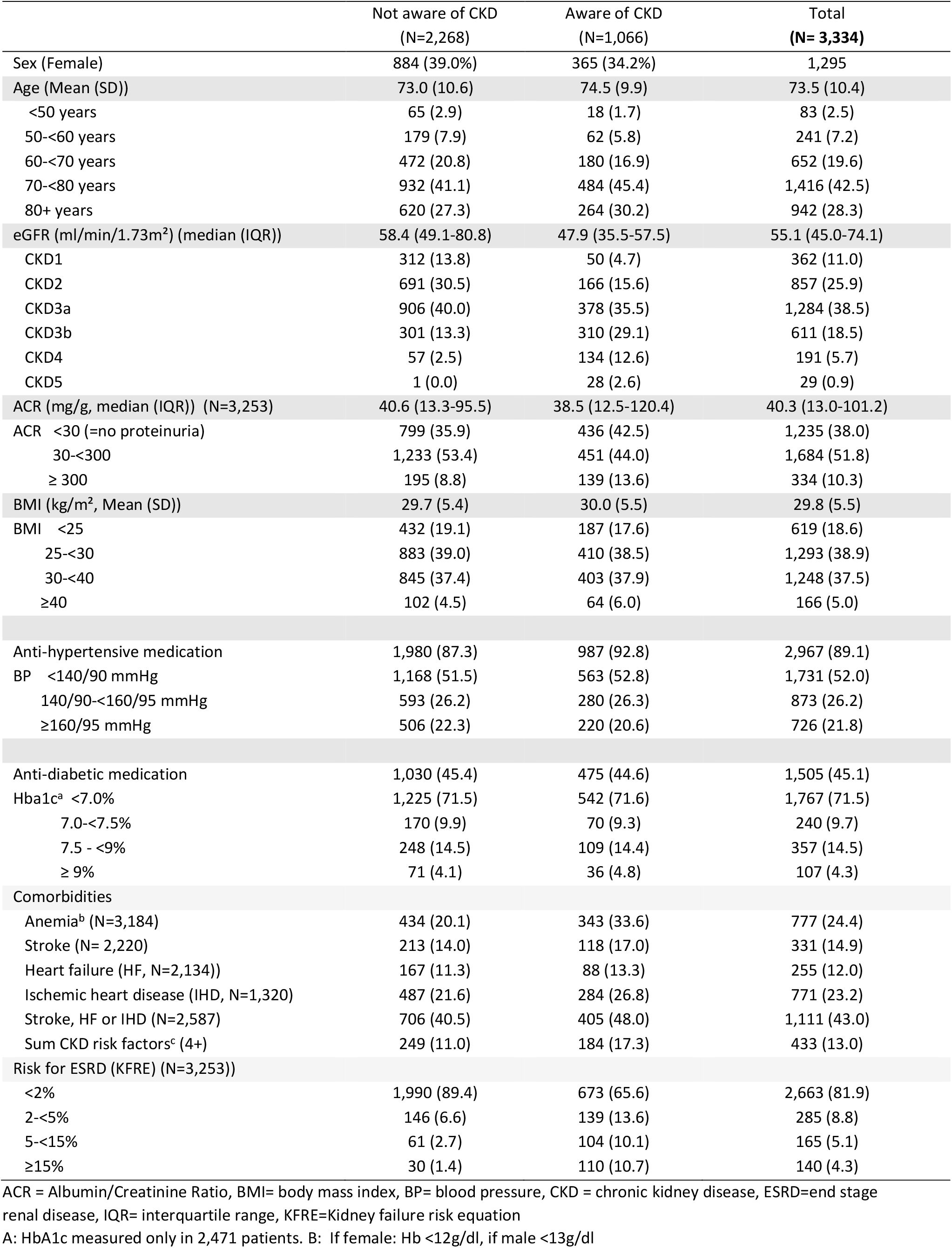
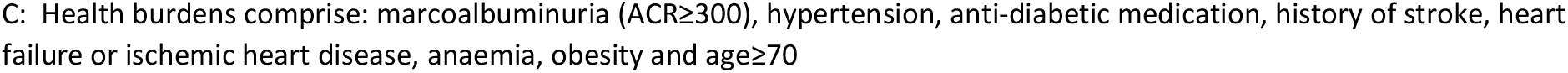
Characteristics of all participants and stratified by awareness for chronic kidney disease (CKD): prevalence (N, %), mean and standard deviation (SD) or median and interquartile ranges (IQR). German CKD related cohorts (CORE database), 2010.

|  | Not aware of CKD<br>(N=2,268) | Aware of CKD<br>(N=1,066) | Total<br>(N= 3,334) |
| --- | --- | --- | --- |
| Sex (Female) | 884 (39.0%) | 365 (34.2%) | 1,295 |
| Age (Mean (SD)) | 73.0 (10.6) | 74.5 (9.9) | 73.5 (10.4) |
| <50 years | 65 (2.9) | 18 (1.7) | 83 (2.5) |
| 50-<60 years | 179 (7.9) | 62 (5.8) | 241 (7.2) |
| 60-<70 years | 472 (20.8) | 180 (16.9) | 652 (19.6) |
| 70-<80 years | 932 (41.1) | 484 (45.4) | 1,416 (42.5) |
| 80+ years | 620 (27.3) | 264 (30.2) | 942 (28.3) |
| eGFR (ml/min/1.73m<sup>2</sup>) (median (IQR)) | 58.4 (49.1-80.8) | 47.9 (35.5-57.5) | 55.1 (45.0-74.1) |
| CKD1 | 312 (13.8) | 50 (4.7) | 362 (11.0) |
| CKD2 | 691 (30.5) | 166 (15.6) | 857 (25.9) |
| CKD3a | 906 (40.0) | 378 (35.5) | 1,284 (38.5) |
| CKD3b | 301 (13.3) | 310 (29.1) | 611 (18.5) |
| CKD4 | 57 (2.5) | 134 (12.6) | 191 (5.7) |
| CKD5 | 1 (0.0) | 28 (2.6) | 29 (0.9) |
| ACR (mg/g, median (IQR)) (N=3,253) | 40.6 (13.3-95.5) | 38.5 (12.5-120.4) | 40.3 (13.0-101.2) |
| ACR <30 (=no proteinuria) | 799 (35.9) | 436 (42.5) | 1,235 (38.0) |
| 30-<300 | 1,233 (53.4) | 451 (44.0) | 1,684 (51.8) |
| ≥ 300 | 195 (8.8) | 139 (13.6) | 334 (10.3) |
| BMI (kg/m<sup>2</sup>, Mean (SD)) | 29.7 (5.4) | 30.0 (5.5) | 29.8 (5.5) |
| BMI <25 | 432 (19.1) | 187 (17.6) | 619 (18.6) |
| 25-<30 | 883 (39.0) | 410 (38.5) | 1,293 (38.9) |
| 30-<40 | 845 (37.4) | 403 (37.9) | 1,248 (37.5) |
| ≥40 | 102 (4.5) | 64 (6.0) | 166 (5.0) |
| Anti-hypertensive medication | 1,980 (87.3) | 987 (92.8) | 2,967 (89.1) |
| BP <140/90 mmHg | 1,168 (51.5) | 563 (52.8) | 1,731 (52.0) |
| 140/90-<160/95 mmHg | 593 (26.2) | 280 (26.3) | 873 (26.2) |
| ≥160/95 mmHg | 506 (22.3) | 220 (20.6) | 726 (21.8) |
| Anti-diabetic medication | 1,030 (45.4) | 475 (44.6) | 1,505 (45.1) |
| Hba1c<sup>a</sup> <7.0% | 1,225 (71.5) | 542 (71.6) | 1,767 (71.5) |
| 7.0-<7.5% | 170 (9.9) | 70 (9.3) | 240 (9.7) |
| 7.5 - <9% | 248 (14.5) | 109 (14.4) | 357 (14.5) |
| ≥ 9% | 71 (4.1) | 36 (4.8) | 107 (4.3) |
| Comorbidities |  |  |  |
| Anemia<sup>b</sup> (N=3,184) | 434 (20.1) | 343 (33.6) | 777 (24.4) |
| Stroke (N= 2,220) | 213 (14.0) | 118 (17.0) | 331 (14.9) |
| Heart failure (HF, N=2,134)) | 167 (11.3) | 88 (13.3) | 255 (12.0) |
| Ischemic heart disease (IHD, N=1,320) | 487 (21.6) | 284 (26.8) | 771 (23.2) |
| Stroke, HF or IHD (N=2,587) | 706 (40.5) | 405 (48.0) | 1,111 (43.0) |
| Sum CKD risk factors<sup>c</sup> (4+) | 249 (11.0) | 184 (17.3) | 433 (13.0) |
| Risk for ESRD (KFRE) (N=3,253)) |  |  |  |
| <2% | 1,990 (89.4) | 673 (65.6) | 2,663 (81.9) |
| 2-<5% | 146 (6.6) | 139 (13.6) | 285 (8.8) |
| 5-<15% | 61 (2.7) | 104 (10.1) | 165 (5.1) |
| ≥15% | 30 (1.4) | 110 (10.7) | 140 (4.3) |

ACR = Albumin/Creatinine Ratio, BMI= body mass index, BP= blood pressure, CKD = chronic kidney disease, ESRD=end stage renal disease, IQR= interquartile range, KFRE=Kidney failure risk equation
A: HbA1c measured only in 2,471 patients. B: If female: Hb <12g/dl, if male <13g/dl
C: Health burdens comprise: macroalbuminuria ( $\text{ACR} \geq 300$ ), hypertension, anti-diabetic medication, history of stroke, heart failure or ischemic heart disease, anaemia, obesity and $\text{age} \geq 70$

### Prevalence of unawareness

Compared to men (66% (64%-68%)), women were more often unaware of their CKD (71% (68%-73%)) (Tab. 2). Combining all CKD stages, unawareness decreased in older age from 78% (68%-87%) in patients <50 years to 66% (62%-69%) in patients aged 80 years and older. Women were in all but one age-group (50-59 years) more often unaware than men (Fig. 1).

**Tab 2.**
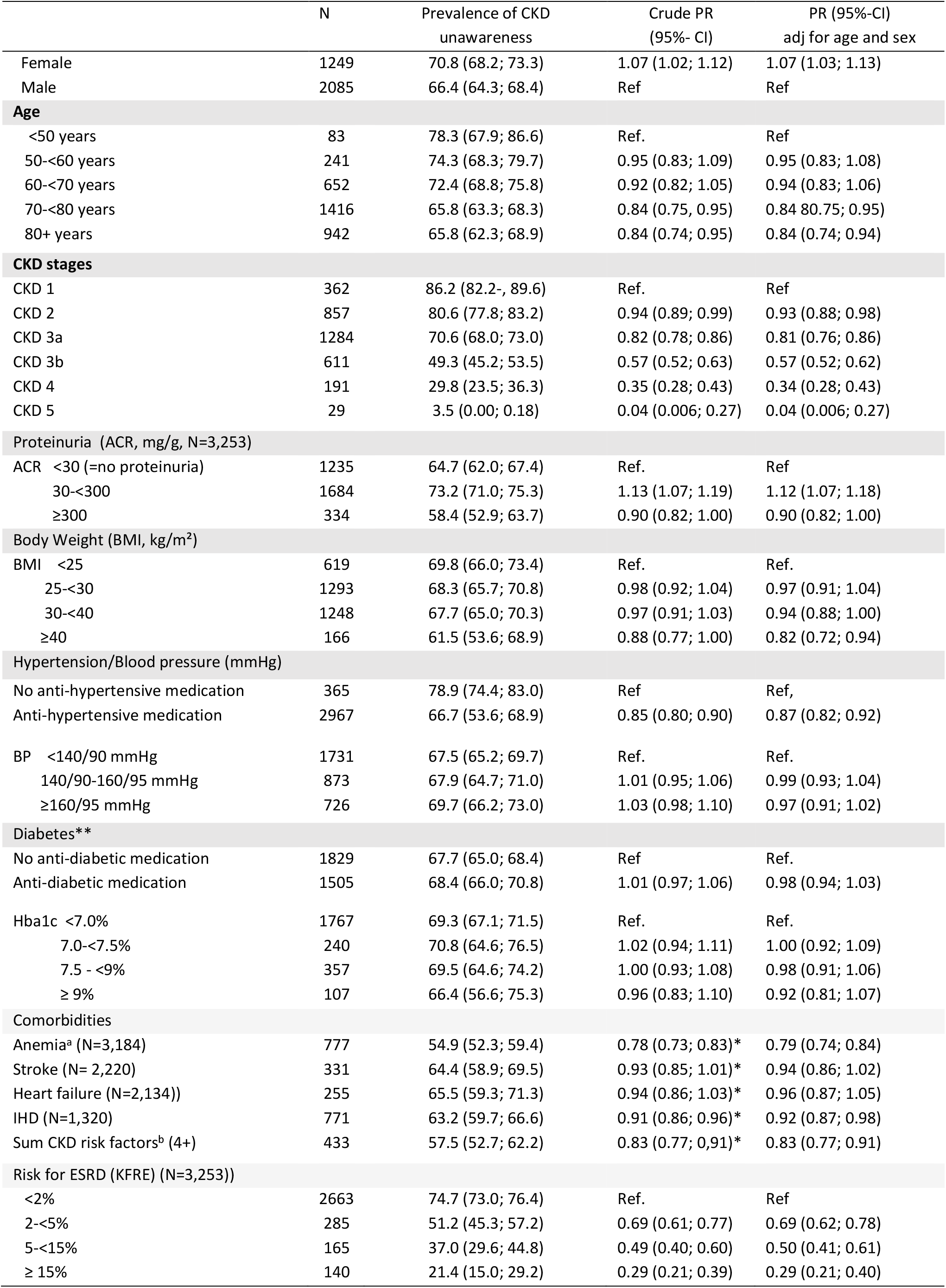
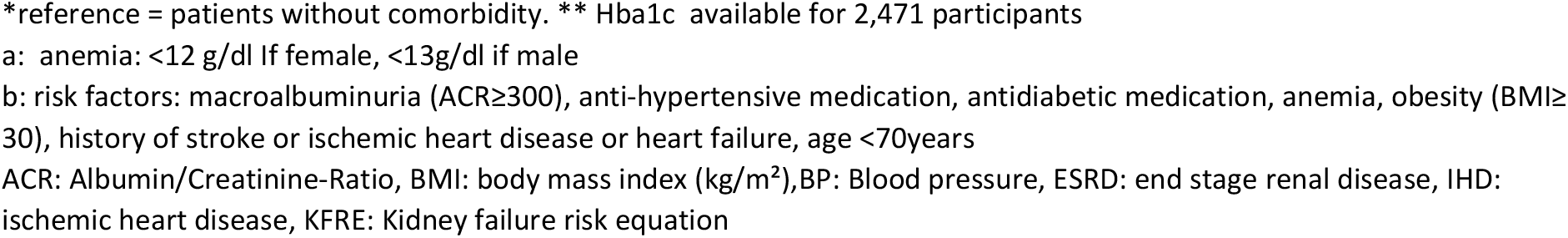
Unawareness for chronic kidney disease (CKD) by patients’ characteristics. Crude and age and sex adjusted prevalence ratios (PR) and 95% confidence intervals (CI) for risk of unawareness. PR for sex adjusted for age, and PR for age adjusted for sex. German CKD cohorts (CORE database), 2010.

|  | N | Prevalence of CKD<br>unawareness | Crude PR<br>(95%- CI) | PR (95%-CI)<br>adj for age and sex |
| --- | --- | --- | --- | --- |
| Female | 1249 | 70.8 (68.2; 73.3) | 1.07 (1.02; 1.12) | 1.07 (1.03; 1.13) |
| Male | 2085 | 66.4 (64.3; 68.4) | Ref | Ref |
| **Age** |  |  |  |  |
| <50 years | 83 | 78.3 (67.9; 86.6) | Ref. | Ref |
| 50-<60 years | 241 | 74.3 (68.3; 79.7) | 0.95 (0.83; 1.09) | 0.95 (0.83; 1.08) |
| 60-<70 years | 652 | 72.4 (68.8; 75.8) | 0.92 (0.82; 1.05) | 0.94 (0.83; 1.06) |
| 70-<80 years | 1416 | 65.8 (63.3; 68.3) | 0.84 (0.75; 0.95) | 0.84 (0.75; 0.95) |
| 80+ years | 942 | 65.8 (62.3; 68.9) | 0.84 (0.74; 0.95) | 0.84 (0.74; 0.94) |
| **CKD stages** |  |  |  |  |
| CKD 1 | 362 | 86.2 (82.2; 89.6) | Ref. | Ref |
| CKD 2 | 857 | 80.6 (77.8; 83.2) | 0.94 (0.89; 0.99) | 0.93 (0.88; 0.98) |
| CKD 3a | 1284 | 70.6 (68.0; 73.0) | 0.82 (0.78; 0.86) | 0.81 (0.76; 0.86) |
| CKD 3b | 611 | 49.3 (45.2; 53.5) | 0.57 (0.52; 0.63) | 0.57 (0.52; 0.62) |
| CKD 4 | 191 | 29.8 (23.5; 36.3) | 0.35 (0.28; 0.43) | 0.34 (0.28; 0.43) |
| CKD 5 | 29 | 3.5 (0.00; 0.18) | 0.04 (0.006; 0.27) | 0.04 (0.006; 0.27) |
| **Proteinuria (ACR, mg/g, N=3,253)** |  |  |  |  |
| ACR <30 (=no proteinuria) | 1235 | 64.7 (62.0; 67.4) | Ref. | Ref |
| 30-<300 | 1684 | 73.2 (71.0; 75.3) | 1.13 (1.07; 1.19) | 1.12 (1.07; 1.18) |
| ≥300 | 334 | 58.4 (52.9; 63.7) | 0.90 (0.82; 1.00) | 0.90 (0.82; 1.00) |
| **Body Weight (BMI, kg/m<sup>2</sup>)** |  |  |  |  |
| BMI <25 | 619 | 69.8 (66.0; 73.4) | Ref. | Ref. |
| 25-<30 | 1293 | 68.3 (65.7; 70.8) | 0.98 (0.92; 1.04) | 0.97 (0.91; 1.04) |
| 30-<40 | 1248 | 67.7 (65.0; 70.3) | 0.97 (0.91; 1.03) | 0.94 (0.88; 1.00) |
| ≥40 | 166 | 61.5 (53.6; 68.9) | 0.88 (0.77; 1.00) | 0.82 (0.72; 0.94) |
| **Hypertension/Blood pressure (mmHg)** |  |  |  |  |
| No anti-hypertensive medication | 365 | 78.9 (74.4; 83.0) | Ref | Ref, |
| Anti-hypertensive medication | 2967 | 66.7 (53.6; 68.9) | 0.85 (0.80; 0.90) | 0.87 (0.82; 0.92) |
| BP <140/90 mmHg | 1731 | 67.5 (65.2; 69.7) | Ref. | Ref. |
| 140/90-160/95 mmHg | 873 | 67.9 (64.7; 71.0) | 1.01 (0.95; 1.06) | 0.99 (0.93; 1.04) |
| ≥160/95 mmHg | 726 | 69.7 (66.2; 73.0) | 1.03 (0.98; 1.10) | 0.97 (0.91; 1.02) |
| **Diabetes\*\*** |  |  |  |  |
| No anti-diabetic medication | 1829 | 67.7 (65.0; 68.4) | Ref | Ref. |
| Anti-diabetic medication | 1505 | 68.4 (66.0; 70.8) | 1.01 (0.97; 1.06) | 0.98 (0.94; 1.03) |
| Hba1c <7.0% | 1767 | 69.3 (67.1; 71.5) | Ref. | Ref. |
| 7.0-<7.5% | 240 | 70.8 (64.6; 76.5) | 1.02 (0.94; 1.11) | 1.00 (0.92; 1.09) |
| 7.5 - <9% | 357 | 69.5 (64.6; 74.2) | 1.00 (0.93; 1.08) | 0.98 (0.91; 1.06) |
| ≥ 9% | 107 | 66.4 (56.6; 75.3) | 0.96 (0.83; 1.10) | 0.92 (0.81; 1.07) |
| **Comorbidities** |  |  |  |  |
| Anemia<sup>a</sup> (N=3,184) | 777 | 54.9 (52.3; 59.4) | 0.78 (0.73; 0.83)* | 0.79 (0.74; 0.84) |
| Stroke (N= 2,220) | 331 | 64.4 (58.9; 69.5) | 0.93 (0.85; 1.01)* | 0.94 (0.86; 1.02) |
| Heart failure (N=2,134)) | 255 | 65.5 (59.3; 71.3) | 0.94 (0.86; 1.03)* | 0.96 (0.87; 1.05) |
| IHD (N=1,320) | 771 | 63.2 (59.7; 66.6) | 0.91 (0.86; 0.96)* | 0.92 (0.87; 0.98) |
| Sum CKD risk factors<sup>b</sup> (4+) | 433 | 57.5 (52.7; 62.2) | 0.83 (0.77; 0.91)* | 0.83 (0.77; 0.91) |
| **Risk for ESRD (KFRE) (N=3,253))** |  |  |  |  |
| <2% | 2663 | 74.7 (73.0; 76.4) | Ref. | Ref |
| 2-<5% | 285 | 51.2 (45.3; 57.2) | 0.69 (0.61; 0.77) | 0.69 (0.62; 0.78) |
| 5-<15% | 165 | 37.0 (29.6; 44.8) | 0.49 (0.40; 0.60) | 0.50 (0.41; 0.61) |
| ≥ 15% | 140 | 21.4 (15.0; 29.2) | 0.29 (0.21; 0.39) | 0.29 (0.21; 0.40) |

\*reference = patients without comorbidity. \*\* Hba1c available for 2,471 participants
a: anemia: <12 g/dl if female, <13g/dl if male
b: risk factors: macroalbuminuria (ACR ≥ 300), anti-hypertensive medication, antidiabetic medication, anemia, obesity (BMI ≥ 30), history of stroke or ischemic heart disease or heart failure, age <70years
ACR: Albumin/Creatinine-Ratio, BMI: body mass index (kg/m<sup>2</sup>), BP: Blood pressure, ESRD: end stage renal disease, IHD: ischemic heart disease, KFRE: Kidney failure risk equation

**Fig. 1.**
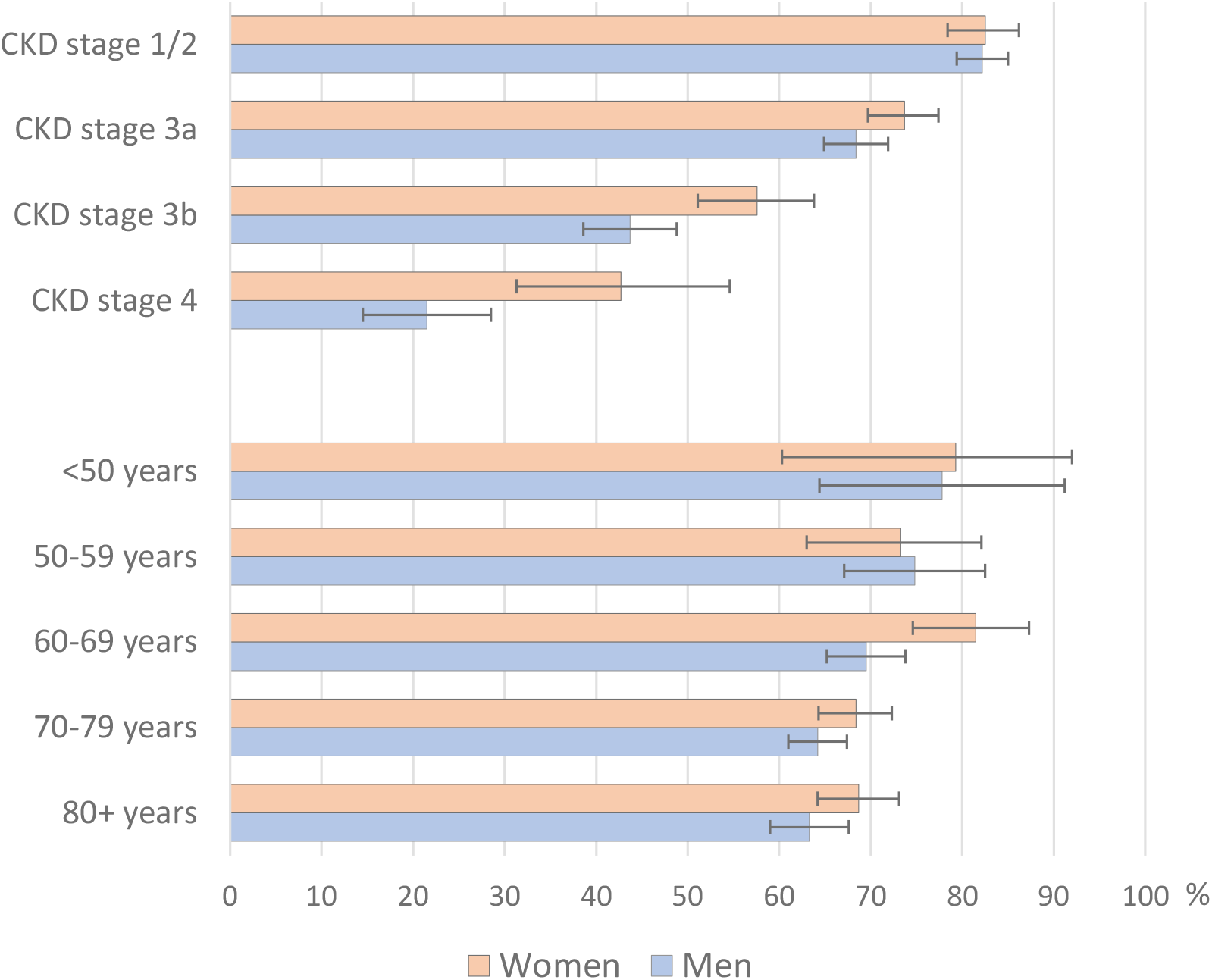
Sex difference in unawareness for chronic kidney disease (CKD) by CKD stage and age groups; prevalence with 95% confidence intervals. German CKD related cohorts (CORE database), 2010.

Unawareness decreased with increasing CKD stage. 82% (80%-84%) of the studies’ participants with CKD stage 1 or 2 were unaware of their CKD. Unawareness amounted to a percentage of 71% (68%-73%) in patients with CKD stage 3a and to 49% (45%-54%) in patients with CKD stage 3b. In patients with CKD stage 4, one out of three patients was unaware of the condition.

Even in patients with CKD-related comorbidities CKD unawareness was high. About two-thirds of patients with a history of cardiovascular diseases, with anti-hypertensive or anti-diabetic medication, with high blood pressure or inadequately controlled HbA1c (>7.5%) did not know about their CKD. Unawareness was lower in patients with macroalbuminuria (58% (53%-64%)), anemia (55% (52%-59%)) and in patients with at least four conditions unfavorable for CKD (58% (53%-62%)) (Fig. 1). Unawareness decreased with increasing risk for ESRD within 5 years. But, still 21% (15%-29%) of the patients with an ESRD risk ≥ 15% did not know about their disease that might lead to renal replacement therapy within the next five years.

### Prevalence of unawareness in regard to CKD stages

*S*tratified according to CKD stage, unawareness decreased with age in CKD stages 1-3a, but increased in later age in later CKD (Tab. 3). While in early CKD, no sex difference in unawareness was visible, women were more often unaware of a CKD in higher CKD stages than men. Eventually, in CKD stage 4, unawareness in women was twice as high than in men (43% (33%-55%) vs. 22% (15%-30%)) (Fig. 1).

**Tab. 3.** Unawareness for chronic kidney disease (CKD) (95%-confidence interval) by demographic and clinical parameters, stratified by CKD stage (N=3,305). German CKD related cohorts (CORE database), 2010.

|  | CKD stage 1/2 | CKD stage 3a | CKD stage 3b | CKD stage 4 |
| --- | --- | --- | --- | --- |
| **Age (yrs)** |  |  |  |  |
| <50 | 87.5 (77.6; 94.1) | 50.0 (0.07; 0.93) | - | - |
| 50-59 | 81.9 (75.5; 87.3) | 76.3 (59.8; 88.6) | 21.4 (0.05; 50.1) | 22.2 (2.8; 60.0) |
| 60-69 | 83.8 (79.5; 87.7) | 70.3 (63.6; 76.4) | 43.1 (30.9; 56.0) | 23.1 (9.0; 43.7) |
| 70-79 | 82.3 (78.4; 85.7) | 69.0 (65.2; 72.7) | 45.8 (39.7; 52.1) | 25.6 (16.4; 36.8) |
| 80+ | 77.6 (70.7; 83.6) | 72.6 (68.1; 76.8) | 56.2 (50.5; 62.3) | 39.2 (28.0; 51.2) |
| **BMI (kg/m, N=6,518)** |  |  |  |  |
| <25 | 86.9 (81.7; 91.0) | 70.5 (64.3; 76.1) | 54.9 (45.2; 64.3) | 19.4 (7.5; 37.5) |
| 25-<30 | 83.3 (79.4; 86.8) | 72.4 (68.4; 76.1) | 46.9 (40.6; 53.2) | 34.3 (23.2; 46.9) |
| 30-<40 | 80.9 (77.2; 84.3) | 69.1 (64.5; 73.4) | 50.2 (43.4; 57.0) | 32.1 (22.2; 43.4) |
| ≥40 | 71.1 (59.5; 80.9) | 64.3 (50.4; 76.6) | 40.9 (20.7; 63.7) | 18.2 (2.3; 51.8) |
| **Anti-hypertensive medication** |  |  |  |  |
| No | 85.4 (79.6; 90.1) | 79.1 (71.2; 85.6) | 59.3 (38.8; 77.6) | 20.0 (2.5; 55.6) |
| Yes | 81.7 (79.2; 84.0) | 69.7 (66.9; 72.3) | 48.8 (44.7; 52.9) | 30.4 (23.8; 37.7) |
| **Blood pressure (mmHg)** |  |  |  |  |
| <140/90 | 83.8 (80.4; 86.7) | 71.0 (67.4; 74.3) | 49.3 (43.8; 54.7) | 31.1 (22.9; 40.2) |
| 140/90-160/95 | 80.6 (76.2; 85.0) | 72.2 (67.2; 77.0) | 47.7 (39.7; 55.9) | 28.6 (16.6; 43.3) |
| >160/95 | 81.4 (76.8; 85.5) | 67.3 (61.2; 73.0) | 51.7 (42.3; 61.1) | 26.1 (10.2; 48.4) |
| **Proteinuria (ACR mg/g)** |  |  |  |  |
| <30 (no proteinuria) | - | 71.3 (68.1; 74.4) | 53.5 (48.0; 59.0) | 37.7 (26.3; 50.2) |
| 30-<300 | 83.1 (80.7; 85.3) | 67.8 (62.6; 72.9) | 47.1 (40.1; 54.2) | 27.3 (17.0; 39.6) |
| ≥300 | 76.6 (68.8; 83.2) | 72.1 (59.9; 82.3) | 38.3 (26.1; 51.8) | 26.1 (14.3; 41.1) |
| **Anti-diabetic medication** |  |  |  |  |
| No | 85.4 (82.1; 88.2) | 71.2 (67.9; 74.4) | 49.6 (44.5; 54.7) | 24.5 (16.4; 34.2) |
| Yes | 79.7 (76.5; 82.7) | 69.6 (65.4; 73.6) | 48.7 (42.0; 55.4) | 35.5 (25.8; 46.1) |
| **Hba1c (% , N=2,461)** |  |  |  |  |
| <7 | 80.1 (76.6; 83.2) | 71.6 (68.2; 74.8) | 56.1 (50.6; 61.5) | 36.6 (26.8; 47.2) |
| 7-<7.5 | 81.3 (72.6; 88.2) | 72.9 (60.9; 82.8) | 47.7 (32.5; 63.3) | 58.8 (32.9; 81.6) |
| 7.5-<9 | 79.5 (72.7; 85.3) | 76.6 (67.5; 84.3) | 41.7 (29.1; 55.1) | 26.3 (9.2; 51.2) |
| 9+ | 74.6 (62.1; 84.7) | 62.5 (40.6; 81.2) | 50.0 (18.2; 81.2) | 40.0 (12.2; 73.8) |
| **Anemia<sup>a</sup> (N=3,155)** |  |  |  |  |
| No | 82.7 (80.3; 85.0) | 71.5 (68.9; 74.8) | 51.5 (46.3; 56.7) | 35.5 (24.9; 47.3) |
| Yes | 76.5 (68.4; 83.3) | 67.5 (61.8; 72.7) | 46.5 (39.8; 53.4) | 25.0 (17.2; 34.3) |
| **Stroke or IHD or heart failure (N=3,289)** |  |  |  |  |
| No | 78.6 (75.0; 81.8) | 72.4 (68.5; 76.0) | 54.0 (47.4; 60.4) | 40.0 (28.0; 52.9) |
| Yes | 81.8 (77.4; 85.8) | 67.4 (62.7; 71.9) | 47.0 (40.6; 53.4) | 26.9 (18.2; 37.1) |
| **Sum of risk factors<sup>b</sup>** |  |  |  |  |
| 0-3 | 83.2 (80.9; 85.4) | 71.2 (68.5; 73.8) | 49.3 (44.9; 53.8) | 28.0 (20.3; 36.7) |
| 4 and more | 73.7 (64.8; 81.4) | 65.2 (56.5; 73.2) | 49.0 (39.1; 59.0) | 33.3 (22.2; 44.7) |

ACR: Albumin/Creatinine Ratio, BMI: body mass index, CKD: chronic kidney disease, IHD: ischemic heart disease.
a: If female: Hb <12g/dl, if male <13g/dl
b: sum of risk factors include: macroalbuminuria (ACR≥300), hypertension, anti-diabetic medication, history of stroke, heart failure or ischemic heart disease, anemia, obesity and age≥70

Unawareness of CKD remained high in later CKD stages (about 50% in CKD stage 3b and 30% in CKD stage 4) in spite of prevalent additional unfavorable health conditions such as hypertension, diabetes or overweight.

### Association between unawareness and comorbidities

Strong obesity (BMI ≥40) (PR= 0.88 (0.77-1.00)), anemia (PR=0.78 (0.73-0.83)), intake of anti-hypertensive medication (PR=0.85 (0.80-0.90)), macroalbuminuria (PR=0.90 (0.82-1.00)) or a history of stroke, heart failure or ischemic heart disease were associated with a lower unawareness in CKD patients (Tab. 2). Microalbuminuria, however, was associated with a higher unawareness for CKD (PR=1.13 (1.07-1.19)).

### Unawareness in diabetic and hypertensive patients

In diabetic patients taking insulin, CKD unawareness was lower (PR=65% (62%-69%) than in patients taking metformin or metformin-combined drugs (PR=79% (67%-82%)) (Tab. 4). This difference increased with the increasing CKD stage. Unawareness in CKD stage 4 was higher in diabetic than in non-diabetic patients. Metformin which is contra-indicated in CKD stage 4 was taken by 11 participants; of these 8 (=73%) were unaware of their CKD. Diabetic patients with less well controlled glucose (HbA1c ≥7,5%) showed a lower unawareness of CKD in all CKD stages compared to diabetic patients reaching the recommended HbA1c of <7.5%.

**Tab 4.**
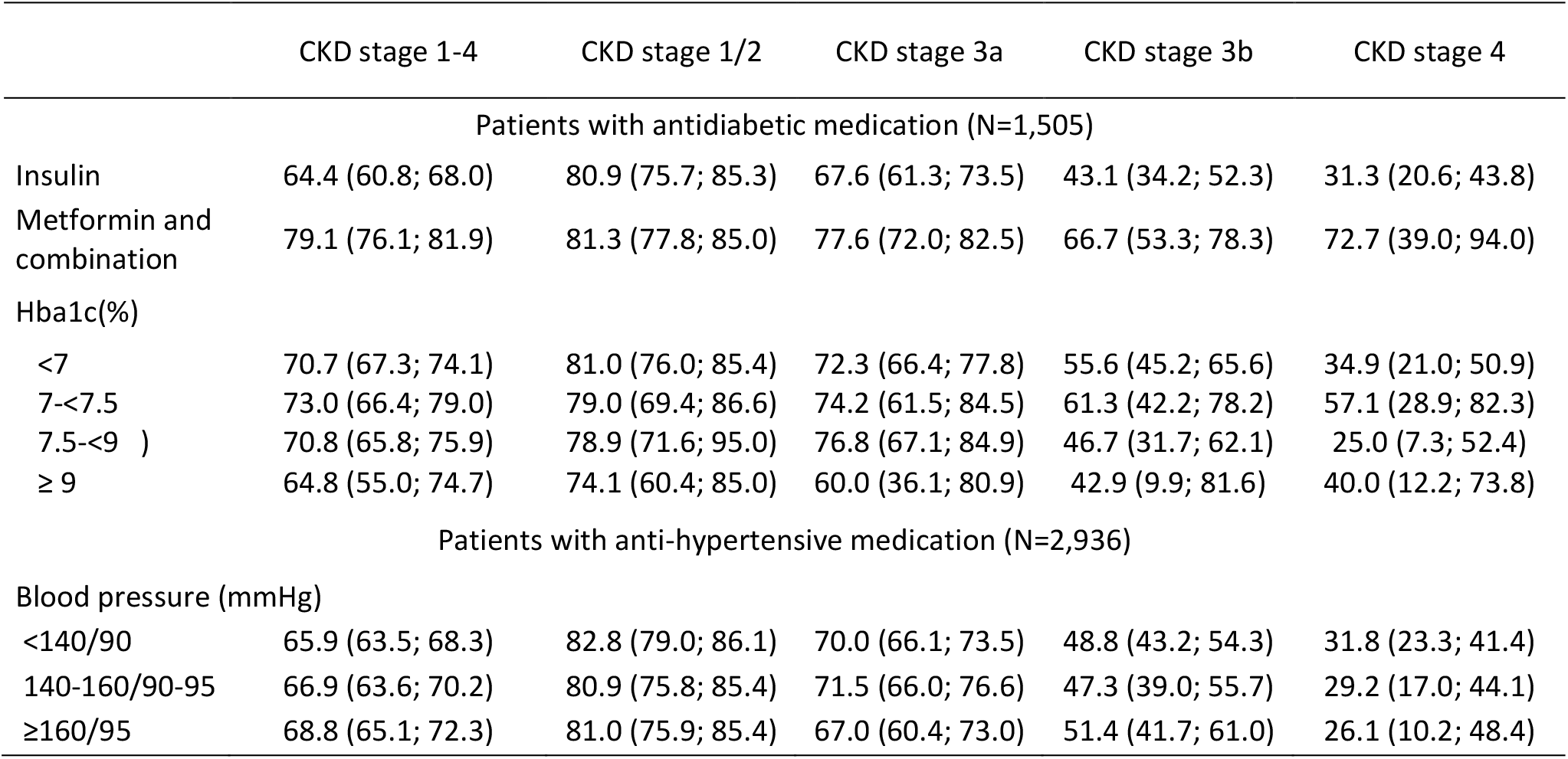
Patients with antidiabetic medication (N=1,505) and anti-hypertensive medication (N=2,936): Prevalence (95% confidence interval) of unawareness for chronic kidney disease (CKD) by type of antidiabetic treatment and glucose control and blood pressure – stratified by CKD stage. German CKD related cohorts (CORE database), 2010.

|  | CKD stage 1-4 | CKD stage 1/2 | CKD stage 3a | CKD stage 3b | CKD stage 4 |
| --- | --- | --- | --- | --- | --- |
| Patients with antidiabetic medication (N=1,505) |  |  |  |  |  |
| Insulin | 64.4 (60.8; 68.0) | 80.9 (75.7; 85.3) | 67.6 (61.3; 73.5) | 43.1 (34.2; 52.3) | 31.3 (20.6; 43.8) |
| Metformin and combination | 79.1 (76.1; 81.9) | 81.3 (77.8; 85.0) | 77.6 (72.0; 82.5) | 66.7 (53.3; 78.3) | 72.7 (39.0; 94.0) |
| Hba1c(%) |  |  |  |  |  |
| <7 | 70.7 (67.3; 74.1) | 81.0 (76.0; 85.4) | 72.3 (66.4; 77.8) | 55.6 (45.2; 65.6) | 34.9 (21.0; 50.9) |
| 7-<7.5 | 73.0 (66.4; 79.0) | 79.0 (69.4; 86.6) | 74.2 (61.5; 84.5) | 61.3 (42.2; 78.2) | 57.1 (28.9; 82.3) |
| 7.5-<9 ) | 70.8 (65.8; 75.9) | 78.9 (71.6; 95.0) | 76.8 (67.1; 84.9) | 46.7 (31.7; 62.1) | 25.0 (7.3; 52.4) |
| ≥ 9 | 64.8 (55.0; 74.7) | 74.1 (60.4; 85.0) | 60.0 (36.1; 80.9) | 42.9 (9.9; 81.6) | 40.0 (12.2; 73.8) |
| Patients with anti-hypertensive medication (N=2,936) |  |  |  |  |  |
| Blood pressure (mmHg) |  |  |  |  |  |
| <140/90 | 65.9 (63.5; 68.3) | 82.8 (79.0; 86.1) | 70.0 (66.1; 73.5) | 48.8 (43.2; 54.3) | 31.8 (23.3; 41.4) |
| 140-160/90-95 | 66.9 (63.6; 70.2) | 80.9 (75.8; 85.4) | 71.5 (66.0; 76.6) | 47.3 (39.0; 55.7) | 29.2 (17.0; 44.1) |
| ≥160/95 | 68.8 (65.1; 72.3) | 81.0 (75.9; 85.4) | 67.0 (60.4; 73.0) | 51.4 (41.7; 61.0) | 26.1 (10.2; 48.4) |

In patients taking anti-hypertensive medication, less well controlled blood pressure was not associated with lower unawareness.

## Discussion

In a population of patients with high prevalence of CKD risk factors, we found a CKD unawareness of 80% in early CKD stages 1 and 2 and still of about 30% in patients with CKD stage 4. Unawareness for CKD was seen even in the elderly or patients with hypertension or diabetes. In these patients, guidelines for treatment and drug prescribing recommend a routine monitoring of renal function. Therefore, the extent of CKD unawareness was unexpected in these subgroups and may reflect a low adherence or knowledge of guidelines. A gender gap in CKD awareness with a higher unawareness in women increased distinctly with increasing stages of CKD.

### Unawareness in patients, physicians and public

Information on CKD awareness was derived from the participants’ answer to the question whether they had ever been told by their physician that they had a renal disease or kidney stones. Therefore, patients’ unawareness could either derive from a lack of understanding the physicians’ information about their CKD or from their physicians’ unawareness of their CKD. 2009, in a report on the prevalence of patients with chronic diseases in general practice, the authoring physicians did not select CKD as one of 20 relevant diseases and conditions (22), although CKD prevalence in this setting is estimated to be about 30% (23). Guidelines on treatment of hypertension or diabetes are much more familiar in general practice than those dealing with CKD (24). It has been shown that interventions to increase CKD knowledge and awareness in primary care physicians can lead to better CKD diagnoses and risk factor management (25).

In about 80% records of patients in general practice (26) and in hospitalized patients due to cardiovascular events (11), a prevalent CKD was not mentioned in their record, less so in patients with diabetes or obese patients. Even patients treated for CKD are often unaware. In patients from a nephrological outpatient clinic, unawareness of CKD stage 1 or 2 was 40%, and in later stages about 12% (27). In UK, 41% of patients with CKD stage 3 that are documented in their GPs CKD registry were unaware of their disease (28).

The high CKD unawareness in patients at higher age (≥70 years) is disturbing. Age-related physiological changes in pharmacodynamics along with decreasing glomerular filtration require renal monitoring and adjustment of prescription and dosing of drugs. About 90% of the older patients in our study reported intake of anti-hypertensive medication. Therefore, they should regularly visit their GP. In our cohort, with increasing age, in CKD stages 3b and 4 unawareness increased also. Physicians seem reluctant to disclose CKD related laboratory findings to their patients. As it is still discussed when to define CKD as ‘disease’ in the elderly (4), physicians do not want to alarm their patients needlessly, when CKD is still not causing any trouble and worry about over-medicalization. However, an EGFR <30ml/min/1.73m^2^ defining CKD stage 4 should be generally regarded as pathologic. On the other hand, if physicians inform their patients about a renal dysfunction, patients may not grasp the meaning and impact it has on their health (29). In an analyses of primary care encounters, CKD was less often discussed than other conditions and information about CKD was given mostly about technical details as laboratory values (14, 30), although better health information facilitates success in patients’ adherence to treatments (13, 31).

### CKD unawareness and additional CKD related risk factors

In our cohort, in 97% of all patients, at least one condition was present which should prompt screening of renal function. But even patients already treated or with diagnostic markers for CKD were often unaware. Albuminuria as marker for CKD seem to trigger renal screening, as unawareness was lower in these patients in our as well as in other cohorts (9). Diabetes, hypertension or cardiovascular diseases were more prevalent in our cohort than in CKD patients in general (32). Diabetic patients are at higher risk for diabetic nephropathy or other renal function disorders. Known diabetes as well as hypertension should trigger monitoring renal function. But, unawareness was about 70% in diabetic or hypertensive patients with CKD stage 3a and 50% in CKD stage 3b. A German study found a similar CKD unawareness in patients with coronary heart disease (11). US studies reported even higher CKD unawareness in patients with diabetes or hypertension (33). In our data, 36% of patients with diabetes and CKD stage 4 were unaware of their CKD. A finding that we find difficult to explain, especially as Metformin, an anti-diabetic drug, is contraindicated then.

### Unawareness and high risk of renal failure

In our cohort, 21% of patients with a ≥15% risk for renal failure within five years were not informed about the critical state of their disease. These patients might lack necessary time for preparation for renal replacement options. In patients from NHANES with CKD stages 3-4, unawareness was even higher among those with a KFRE risk of ≥15% (50%) (34).

### Gender gap in awareness

The gender gap in CKD awareness was unexpected, especially since this gap increased strongly with decreasing renal function. If unawareness for CKD in higher stages can be associated with higher probability of non-treatment or non-adherence to a treatment, women will have a higher risk for CKD-related adverse health outcomes such as cardiovascular diseases, hospitalizations and premature mortality. In 187 CKD participants with CKD from a German population, sex differences in CKD awareness were not visible (10). A recent analysis of NHANES data found a higher CKD unawareness in women compared to men, but only in the Caucasian participants. Sex differences were smaller than in our cohort (35). The reason, why women are more unaware about a potential critical CKD is difficult to explain. Women were similarly affected by comorbidities which should require renal monitoring by their physician. It could have been expected, that they get information about a CKD in the same manner as men. Women have been shown to be more interested in, and are more active seeking, health-related information than men (36). Men, independent of educational attainment, are less engaged in healthy lifestyles than women (37) including less proactive and preventive behavior (38). Sex differences in treatment and disease outcome are also related to physicians’ bias and unconscious attitudes towards female and male patients (39).

### Strengths and limitations

We analyzed a unique large high quality data base on patients with CKD who represent a wide range of relevant comorbidities and ages. Our analyses highlight the extent of CKD unawareness in patients at higher CKD risk in general clinical practice.

However, we do not have information on educational attainment or social status, which might be associated with patient awareness. In German CKD patients, unawareness was not associated with educational attainment (10). As the patients in our cohort took part in a study or register, they might have been more interested in their health than the average patient. Generally, study participants seem to have higher education than the general population (40). Therefore, the prevalence of unawareness in CKD patients in the general population might be underestimated in our analyses.

### Conclusion

CKD is even prevalent as other publicly better known chronic diseases as diabetes or coronary heart disease. But, awareness for CKD falls short. Even in a country with universal and free health coverage, guideline-recommended monitoring of renal function in patients at higher risk for CKD do not reach the practice. Most alarmingly, independent of age, women’s unawareness, especially of later CKD stages, is higher than mens’. Potential explanations do require further, more detailed research in general, and gender specific CKD related patient-physician communication and treatment.

What is already known on this subject?

- Unawareness of a prevalent CKD is large in asymptomatic early stages and is still considerably widespread in later stages of the disease
- Comorbidities such as cardiovascular disease do not lead to a rise in awareness of CKD

What this study adds

- Independent of age, women’s unawareness, especially of later CKD stages, is higher than mens’.
- Guideline recommendations for monitoring renal function do not reach the practice: In a country with universal free health coverage, unawareness is high even in patients at higher risk for CKD.

## Supporting information

see supplementary file

## Data Availability

Data is not available for analyses

## Funding

The CORE-database was by the foundation for preventive medicine of the board of trustees for dialysis and renal transplantation (Stiftung Präventivmedizin des Kuratoriums für Dialyse und Nierentransplantation e. V., KfH).

SS and CS were employees of the foundation for preventive medicine of the board of trustees for dialysis and renal transplantation (Stiftung Präventivmedizin des Kuratoriums für Dialyse und Nierentransplantation e. V., KfH) (no grant number available).

The foundation for preventive medicine of the board of trustees for dialysis and renal transplantation covered travel expenses in course of this research project for SS and CS.

## Acknowledgements

The authors also thank the Advisory Board of the DIACORE-Study: C. Boeger and B. Jung, Regensburg, Germany; CAD-REF-Register: E. Brand and H. Pavenstaedt, Münster, Germany; BIS-Study: E. Schaeffner and N. Ebert, Berlin, Germany; GCKD-Study: K.-U. Eckardt and H. Meiselbach, Berlin and Erlangen, Germany.

## Ethics

CAD-REF: An ethics committee approval was obtained from the Ethics Committee of the Landesärztekammer Westfalen-Lippe and the Medical Faculty of the Westfälische Wilhelms-University Muenster in 2007 (No. 2007-315-f-S).

BIS:The study was approved by the ethics committee of Charite’ University medicine, Berlin in 2008 (No. EA2/009/08)

DIACORE: Ethics committee approval from the universities of Regensburg and Mannheim was obtained in 2007 (No. 06/139) and 2012 (Nr 2012-248N-MA).

## Literature

1. Baumeister SE, Boger CA, Kramer BK, Doring A, Eheberg D, Fischer B, et al. Effect of chronic kidney disease and comorbid conditions on health care costs: A 10-year observational study in a general population. Am J Nephrol. 2010;31(3):222–9.

2. Bruck K, Stel VS, Gambaro G, Hallan S, Volzke H, Arnlov J, et al. CKD Prevalence Varies across the European General Population. J Am Soc Nephrol. 2016;27(7):2135–47.

3. Ng JK, Li PK. Chronic kidney disease epidemic: How do we deal with it? Nephrology (Carlton). 2018;23 Suppl 4:116–20.

4. Delanaye P, Jager KJ, Bokenkamp A, Christensson A, Dubourg L, Eriksen BO, et al. CKD: A Call for an Age-Adapted Definition. J Am Soc Nephrol. 2019;30(10):1785–805.

5. Bretschneider J, Kuhnert R, Hapke U. [Depressive symptoms in adults in Germany]. J Health Monitoring. 2017;2(3):81–8.

6. Gößwald A, Schienkiewitz A, Nowossadeck E, Busch MA. [Prevalence of myocardial infarction and coronary heart disease in adults aged 40-79 years in Germany: results of the German Health Interview and Examination Survey for Adults (DEGS1)]. Bundesgesundheitsblatt Gesundheitsforschung Gesundheitsschutz. 2013;56(5-6):650–5.

7. Heidemann C, Scheidt-Nave C. [Prevalence, incidence and mortality of diabetes mellitus in adults in Germany]. J Health Monitoring. 2017;2 (3):105–29.

8. Hsiao LL. Raising awareness, screening and prevention of chronic kidney disease: It takes more than a village. Nephrology (Carlton). 2018;23 Suppl 4:107–11.

9. Tuot DS, Plantinga LC, Hsu CY, Jordan R, Burrows NR, Hedgeman E, et al. Chronic kidney disease awareness among individuals with clinical markers of kidney dysfunction. Clin J Am Soc Nephrol. 2011;6(8):1838–44.

10. Girndt M, Trocchi P, Scheidt-Nave C, Markau S, Stang A. The Prevalence of Renal Failure. Results from the German Health Interview and Examination Survey for Adults, 2008-2011 (DEGS1). Dtsch Arztebl Int. 2016;113(6):85–91.

11. Wagner M, Wanner C, Schich M, Kotseva K, Wood D, Hartmann K, et al. Patient’s and physician’s awareness of kidney disease in coronary heart disease patients - a cross-sectional analysis of the German subset of the EUROASPIRE IV survey. BMC Nephrol. 2017;18(1):321.

12. Welch JL, Bartlett Ellis RJ, Perkins SM, Johnson CS, Zimmerman LM, Russell CL, et al. Knowledge and Awareness Among Patients with Chronic Kidney Disease Stage 3. Nephrol Nurs J. 2016;43(6):513–9.

13. Greene J, Hibbard JH. Why does patient activation matter? An examination of the relationships between patient activation and health-related outcomes. J Gen Intern Med. 2012;27(5):520–6.

14. Greer RC, Cooper LA, Crews DC, Powe NR, Boulware LE. Quality of patient-physician discussions about CKD in primary care: a cross-sectional study. Am J Kidney Dis. 2011;57(4):583–91.

15. Dorhofer L, Lammert A, Krane V, Gorski M, Banas B, Wanner C, et al. Study design of DIACORE (DIAbetes COhoRtE) - a cohort study of patients with diabetes mellitus type 2. BMC Med Genet. 2013;14:25.

16. Brand E, Pavenstädt H, Schmieder RE, Engelbertz C, Fobker M, Pinnschmidt HO, et al. The Coronary Artery Disease and Renal Failure (CAD-REF) registry: trial design, methods, and aims. Am Heart J. 2013;166(3):449–56.

17. Schaeffner ES, Ebert N, Delanaye P, Frei U, Gaedeke J, Jakob O, et al. Two novel equations to estimate kidney function in persons aged 70 years or older. Ann Intern Med. 2012;157(7):471–81.

18. Stevens PE, Levin A. Evaluation and management of chronic kidney disease: synopsis of the kidney disease: improving global outcomes 2012 clinical practice guideline. Ann Intern Med. 2013;158(11):825–30.

19. Trocchi P, Girndt M, Scheidt-Nave C, Markau S, Stang A. Impact of the estimation equation for GFR on population-based prevalence estimates of kidney dysfunction. BMC Nephrol. 2017;18(1):341.

20. Tangri N, Grams ME, Levey AS, Coresh J, Appel LJ, Astor BC, et al. Multinational Assessment of Accuracy of Equations for Predicting Risk of Kidney Failure: A Meta-analysis. JAMA. 2016;315(2):164–74.

21. Hernan MA. The C-Word: Scientific Euphemisms Do Not Improve Causal Inference From Observational Data. Am J Public Health. 2018;108(5):616–9.

22. Edigi G, Schelp H. Prävalenz chronischer Krankheiten und Qualitätsindikatoren in einer Bremer Hausarztpraxis. Z Allg Med. 2009;85 187–95.

23. Gergei I, Klotsche J, Woitas R, Pieper L, Wittchen H, Krämer B, et al. Chronic kidney disease in primary care in Germany. J Public Health 2017;25:223–30.

24. Plantinga LC, Tuot DS, Powe NR. Awareness of chronic kidney disease among patients and providers. Adv Chronic Kidney Dis. 2010;17(3):225–36.

25. Harvey G, Oliver K, Humphreys J, Rothwell K, Hegarty J. Improving the identification and management of chronic kidney disease in primary care: lessons from a staged improvement collaborative. Int J Qual Health Care. 2015;27(1):10–6.

26. Kitsos A, Peterson GM, Jose MD, Khanam MA, Castelino RL, Radford JC. Variation in Documenting Diagnosable Chronic Kidney Disease in General Medical Practice: Implications for Quality Improvement and Research. J Prim Care Community Health. 2019;10:2150132719833298.

27. Devraj R, Borrego ME, Vilay AM, Pailden J, Horowitz B. Awareness, self-management behaviors, health literacy and kidney function relationships in specialty practice. World J Nephrol. 2018;7(1):41–50.

28. McIntyre NJ, Fluck R, McIntyre C, Taal M. Treatment needs and diagnosis awareness in primary care patients with chronic kidney disease. Br J Gen Pract. 2012;62(597):e227–32.

29. Greer RC, Liu Y, Cavanaugh K, Diamantidis CJ, Estrella MM, Sperati CJ, et al. Primary Care Physicians’ Perceived Barriers to Nephrology Referral and Co-management of Patients with CKD: a Qualitative Study. J Gen Intern Med. 2019;34(7):1228–35.

30. Greer RC, Crews DC, Boulware LE. Challenges perceived by primary care providers to educating patients about chronic kidney disease. J Ren Care. 2012;38(4):174–81.

31. Consortium E. EMPAHTiE - Empowering Patients in the management of chronic diseases. European Commission; 2014. Contract No.: Contract Number 2013 62 01.

32. Collins AJ, Gilbertson DT, Snyder JJ, Chen SC, Foley RN. Chronic kidney disease awareness, screening and prevention: rationale for the design of a public education program. Nephrology (Carlton). 2010;15 Suppl 2:37–42.

33. Obadan NO, Walker RJ, Egede LE. Independent correlates of chronic kidney disease awareness among adults with type 2 diabetes. J Diabetes Complications. 2017;31(6):988–91.

34. Chu CD, McCulloch CE, Banerjee T, Pavkov ME, Burrows NR, Gillespie BW, et al. CKD Awareness Among US Adults by Future Risk of Kidney Failure. Am J Kidney Dis. 2020;76(2):174–83.

35. Hodlmoser S, Winkelmayer WC, Zee J, Pecoits-Filho R, Pisoni RL, Port FK, et al. Sex differences in chronic kidney disease awareness among US adults, 1999 to 2018. PLoS One. 2020;15(12):e0243431.

36. Ek S. Gender differences in health information behaviour: a Finnish population-based survey. Health Promot Int. 2015;30(3):736–45.

37. Dawson KA, Schneider MA, Fletcher PC, Bryden PJ. Examining gender differences in the health behaviors of Canadian university students. J R Soc Promot Health. 2007;127(1):38–44.

38. Avdic D, Hägglund P, Lindahl B, Johansson P. Sex differences in sickness absence and the morbidity-mortality paradox: a longitudinal study using Swedish administrative registers. BMJ Open. 2019;9(8):e024098.

39. Chapman EN, Kaatz A, Carnes M. Physicians and implicit bias: how doctors may unwittingly perpetuate health care disparities. J Gen Intern Med. 2013;28(11):1504–10.

40. Enzenbach C, Wicklein B, Wirkner K, Loeffler M. Evaluating selection bias in a population-based cohort study with low baseline participation: the LIFE-Adult-Study. BMC Med Res Methodol. 2019;19(1):135.

