## Supplementary material for "Unawareness for chronic kidney disease is high in all stages, age groups and comorbidities – and higher in women than in men": see supplementary file

### Characteristics of the German CORE database

#### Background

The CORE database was launched in 2009 from the KfH-Foundation for preventive medicine of the board of trustees for dialysis and renal transplantation (KfH-Stiftung Präventivmedizin des Kuratoriums für Dialyse und Nierentransplantation e. V.).

The foundation provided scientific funding for German-wide research projects on questions on early diagnosis, disease-progression indicators and optimization of therapy of chronic kidney disease in all KDIGO-stages and patients affected by CKD of all ages. Since 2009, firstly five cohort studies with CKD patients and secondly studies on healthcare research were financed. Four CKD studies and one registry collected data on study-specific questions and measures, but provided a standardized set of core variables including life style questions, questions on family history and underlying system diseases, health care aspects, kidney and not-kidney related organ specific symptoms, diagnose-related groups, operation and procedure keys and hospitalizations. These core variables were transferred into the CORE database according to a standardized protocol. All laboratory data was analyzed in one central laboratory. Follow-up time for these studies was up to 10 years with a mean of 6-7 years.

Contributing studies/registries are:

- GCKD (German Chronic Kidney Disease Study): Included CKD patients in nephrological care to describe progress and consequences of CKD. Biomarker and genetic factors related to CKD which can be used for prediction of the pace of CKD progress and incidence of cardiovascular complications.
- DIACORE (DIAbetes COhoRtE): analyses of life-style and genes in order to identify parameters that trigger the emergence of adverse complications in some patients but not in the others. Included in DIACORE were patients with diabetes mellitus Type 2.
- CAD-REF (Coronary-Artery-Disease- Renal Failure): a German-wide registry with either patients with a coronary artery disease. Patients with and without a CKD were included. The registry evaluated diagnostic markers and genetic factors using an extensive biobank for analyses of coronary diseases and (incidence of) renal insufficiency.
- BIS (Berlin Initiative Study): described the health status of elderly (70 years and older) with especially focusing on chronic kidney disease. Participants were members of a statutory health insurance in different medical practices in Berlin, representative for the general older population. Patient interviews included questions on life style, comorbidities and medications. The study set up focused on the development of an algorithm to determine the renal function in older aged patients (BIS2-equation).
- 4C (Cardiovascular comorbidity in Children with Chronic kidney disease): analysed severity, progression and factors leading to incident cardiovascular diseases in children with CKD. Children with CKD 3b-5 from 12 European countries aged 6-18 were included.

**Content of the CORE database**

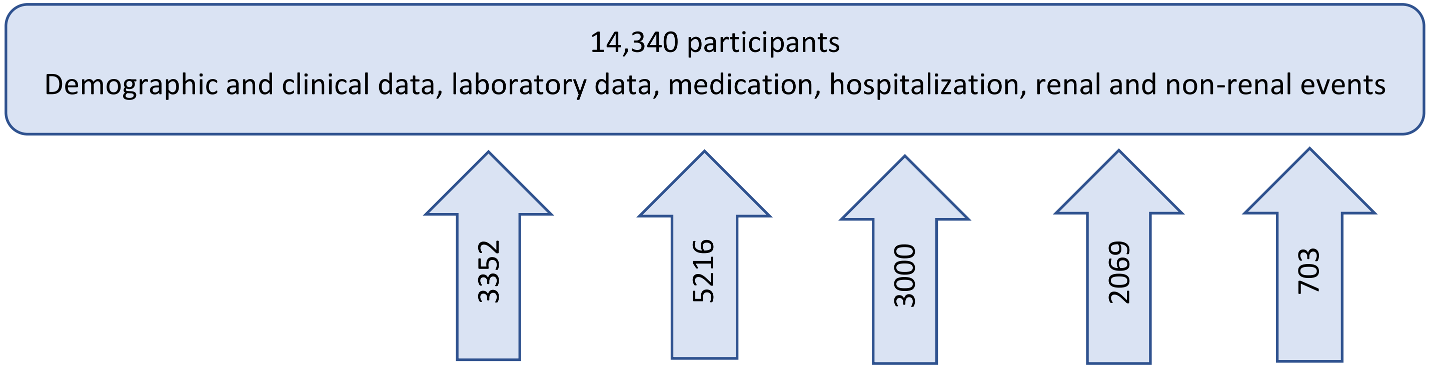


**and characteristics of participants**

| | GCKD | CADRef | DIACORE | BIS | 4C |
|---|---|---|---|---|---|
| ≥60 years | 62% | 76% | 79% | 100% | 0% |
| Unknown CKD | 1% | 59% | 56% | 53% | 1% |
| CKD stage ½ | 21% | 14% | 28% | 21% | 22% |
| CKD 3-5 | 78% | 23% | 20% | 38% | 74% |
| Diabetes[1] | 38% | 32% | 88% | 20% | 0.4% |
| Hypertension[2] | 89% | 83% | 81% | 78% | 37% |

1: Intake of anti-diabetic medication

2: Intake of anti-hypertensive medication
